# The association between thrombocytosis and subtype of lung cancer: a systematic review and meta-analysis

**DOI:** 10.1101/2020.06.08.20125187

**Authors:** Melissa Barlow, Willie Hamilton, Obioha Chukwunyere Ukoumunne, Sarah Elizabeth Rose Bailey

## Abstract

**Background:** Thrombocytosis is associated with poor lung cancer prognosis and has recently been identified as having a high predictive value in lung cancer detection. Lung cancer has multiple histological and genetic subtypes and it is not known whether platelet levels differ across subtypes.

**Methods:** *PubMed* and *Embase* were systematically searched for studies that reported pre-treatment platelet count, as either averages or proportion of patients with thrombocytosis, by histological subtype of lung cancer. Suitable studies were synthesised in meta-analyses; subgroup analyses examined for differences across subtypes.

**Results:** The prevalence of pre-treatment thrombocytosis across all lung cancer patients was 27% (95% CI: 17 to 37%). By subtype, this was 22% (95% CI: 7 to 41%) for adenocarcinoma (ADC), 28% (95% CI: 15 to 43%) for squamous cell carcinoma (SCC), 36% (95% CI: 13 to 62%) for large cell carcinoma, and 30% (95% CI: 8 to 58%) for small cell lung cancer (SCLC). The pooled mean platelet count for lung cancer patients was 289×10^9^ /L (95% CI: 268 to 311). By subtype, this was 282×10^9^ /L (95% CI: 259 to 306) for ADC, 297×10^9^ /L (95% CI: 238 to 356) for SCC, 290×10^9^ /L (95% CI: 176 to 404) for LCC, and 293×10^9^ /L (95% CI: 244 to 342) for SCLC. There was no difference in thrombocytosis prevalence (p=0.76) or mean platelet count (p=0.96) across the subtypes.

**Conclusion:** We report no evidence of differences in platelet levels across the major subtypes of lung cancer. A high platelet level is likely to be generic across all lung cancer subtypes.

**KEY MESSAGES:** *What is the key question?:* Which (if any) subtype(s) of lung cancer are more associated with thrombocytosis?

*What is the bottom line?:* This can facilitate lung cancer diagnostics and provide insights into the biological mechanism between lung cancer and thrombocytosis.

*Why read on?:* This is the first systematic review to compare platelet count across different subtypes of lung cancer, combining data from 9,891 patients across 38 studies.

## INTRODUCTION

Lung cancer is the leading cause of cancer death worldwide. Patients often present at an advanced stage and as a result, the five-year survival rate for lung cancer in the UK is only 16.2%. ^1^ As such, research has focused on identifying biomarkers that can help identify lung cancer at earlier stages and therefore improve survival rates. Thrombocytosis (a peripheral platelet level of >400 × 10^9^ /L of blood) has been long recognised as a marker of poor prognosis in lung cancer patients. ^2^ Recently thrombocytosis has been identified as having a particularly high predictive value for the identification of undiagnosed lung cancer, ^3^ thus making it a promising biomarker in lung cancer diagnostics. However, lung cancer is heterogeneous, with multiple histological and genetic subtypes, and these subtypes each have distinct pathologies, treatment options, and prognoses. Histological lung cancer subtypes are broadly divided into non-small cell lung cancer (NSCLC), which represents 80-85% of all lung cancers, and small cell lung cancer (SCLC) which represents 15-20%. NSCLC is further divided into adenocarcinoma (ADC), squamous cell carcinoma (SCC) and large cell carcinoma (LCC). There are several less common histological subtypes, including adenosquamous carcinoma, though distinguishing between these subtypes can be difficult. In addition to the diversity of the histological subtypes, lung cancers can also be distinguished by their genetic subtype. Most lung cancer genetic mutations are somatic, acquired through environmental factors such as tobacco smoking, and include driver mutations in genes such as *EGFR, KRAS* and *TP53* and rearrangements in the *ALK* gene. Currently, it is not known whether the platelet level differs across subtypes of lung cancer, and therefore whether thrombocytosis is a generic feature of lung cancer, or specific to some subtypes. The answer may facilitate improved lung cancer detection while also providing insights into the biological mechanism between lung cancer and thrombocytosis.

The aim of this systematic review and meta-analysis is to compare lung cancer patients’ pre-treatment platelet counts across lung cancer subtypes to identify which (if any) subtype(s) are more associated with thrombocytosis.

## METHODS

### Search Strategy

This systematic review and meta-analysis followed MOOSE ^4^ and PRISMA-P ^5^ guidelines. We identified relevant studies that reported the platelet count, either as an average or as the proportion of patients with thrombocytosis, by histological subtype of lung cancer. We searched *PubMed* and *Embase* on the 14^th^ January 2020 using search terms related to lung cancer *and thrombocytosis or* platelet count without a date limit (full search terms in Appendix 1). Inclusion criteria for the meta-analysis were: participants were diagnosed with a specific histological or genetic subtype of lung cancer; participants were aged 18 years or over; reported mean platelet count or diagnosis of thrombocytosis before treatments such as surgery, chemotherapy or radiotherapy (as these can affect platelet counts); studies must have reported the mean platelet count or prevalence of thrombocytosis by subtype of lung cancer. Studies were independently screened by two reviewers; conflicts were resolved by discussion. For studies that reported the same cohorts of participants, the study with the largest sample size was selected for the meta-analysis. Studies that reported median platelet count or did not use a thrombocytosis threshold of 400 × 10^9^ /L were excluded from the meta-analysis and were discussed narratively.

### Quality Assessment

Study quality was assessed using the Newcastle-Ottawa Scale ^6^ independently by three reviewers. Scoring was based on three domains: selection, comparability and exposure/outcome for a maximum of 9 points, based on the aims reported in the study rather than the aims of this review. The mean was taken of the three reviewers’ scores and studies were judged as ‘good’, ‘fair’, or ‘poor’ quality based on the Newcastle-Ottawa Scale thresholds. ^6^ As the main extracted data for this analysis were the histological diagnoses and average platelet count (or thrombocytosis proportions), rather than the outcome of the studies, studies rated as ‘poor’ were not excluded from the analyses; however, a sensitivity analysis was conducted to investigate the potential influence of study quality on the findings.

### Data extraction

The following data were extracted independently by two reviewers and cross-referenced for discrepancies: country of study, age of participants, sex ratio, smoking status, number of patients with each histological subtype, and average platelet count and/or proportion of patients with thrombocytosis. Study authors were contacted where data were missing or to confirm whether the inclusion criteria was met. Reminder emails were sent at two monthly intervals if no reply was received.

### Analysis

Analyses were carried out using STATA version 16. Random effects meta-analysis was used to pool the mean platelet count and the proportion of patients with thrombocytosis, using the *meta set* and *metaprop* commands, respectively. The restricted maximum likelihood estimation method was used to pool the mean platelet counts, and when pooling proportions the Freeman-Tukey double arcsine transformation was used to stabilise the variances. Findings were graphically displayed using forest plots. A subgroup analysis compared pooled mean platelet counts and the pooled proportions of patients with thrombocytosis across histological subtypes of lung cancer. The I^2^ statistic quantified heterogeneity. Sensitivity analyses explored potential sources of heterogeneity: first examining only advanced stage cancers, second removing the papers scoring ‘poor’ on the Newcastle-Ottawa Scale, ^6^ and third removing studies that had a small sample size (less than 50 participants) for each histological subtype.

## RESULTS

### Data retrieval and study quality

6,118 studies were returned from the search, including 1,352 duplicates. Abstracts of the remaining 4,766 studies were screened, resulting in 82 studies eligible for full-text screening. Thirty-eight studies met the criteria for the narrative synthesis with a total of twenty studies that fulfilled the inclusion criteria for meta-analysis (Figure 1). Eighteen studies were assessed as ‘good’ quality, five as ‘fair’, and fifteen as ‘poor’ according to the Newcastle-Ottawa Scale thresholds. ^6^

**Figure 1:**
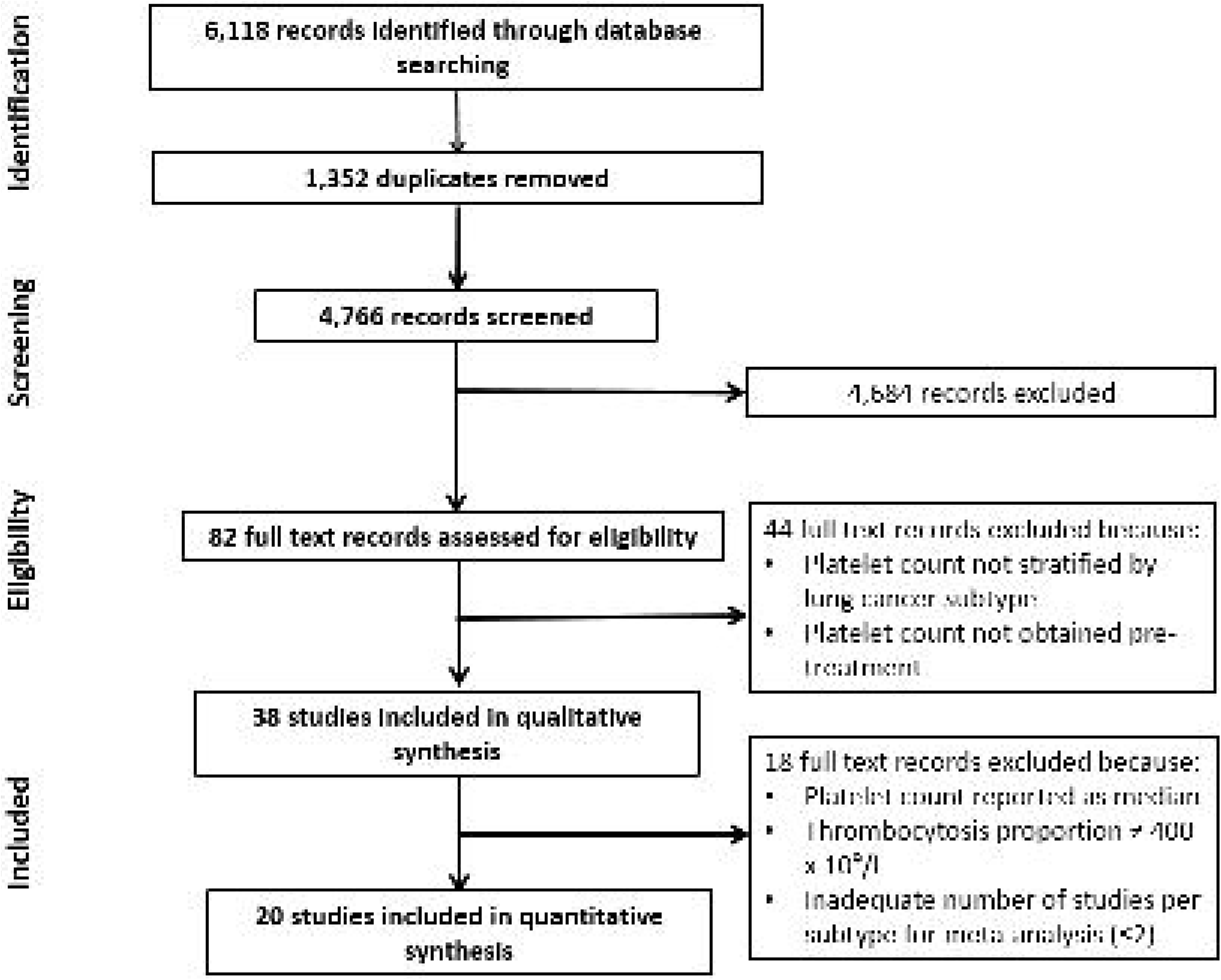
A flow diagram of the study selection process in accordance to PRISMA-P^5^ guidelines.

### Study characteristics

There was a total of 2,974 patients across 20 different studies included in the meta-analyses (Table 1). This consisted of 1,894 patients with ADC, 777 patients with SCC, 55 patients with LCC, and 248 patients with SCLC. Study countries included Bulgaria, Canada, China, Denmark, Greece, India, Japan, Korea, Poland, and Turkey. The ages ranged from 20 to 92 with a mean age of 58 years and median age of 63 years. Three-quarters (73.5%) of participants were male. Sample size ranged from 17 to 347 with a median sample size of 61. Twelve studies reported mean platelet counts and ten studies reported the proportion of patients with thrombocytosis (defined as a platelet level of over 400 × 10^9^ /L); two studies reported both. Seven studies were assessed as ‘good’ quality, four as ‘fair’, and nine as ‘poor’.

**Table 1:** **Characteristics of studies included in meta-analyses thrombocytosis proportions ^7-16^ and mean platelet count. ^12 13 17-26^ ADC, adenocarcinoma; ED, extensive disease; LCC, large cell carcinoma; LD, limited disease; NR – not reported; SCC, squamous cell carcinoma; SCLC, small cell lung cancer.**

| Percentage of patients with thrombocytosis |  |  |  |  |  |  |  |  |
| --- | --- | --- | --- | --- | --- | --- | --- | --- |
| Study | Study Design | Country | Males (%) | Mean age, years (SD) | Smoking history (%) | Clinical stage | Histological subtype, n | Thrombocytosis (%) |
| Alexandrakis (2002)<sup>7</sup> | Case-Control | Greece | 72% | 66 (NR) | NR | NR | ADC, 20<br>SCC, 8<br>LCC, 2<br>SCLC, 11 | 50%<br>50%<br>100%<br>64% |
| Davidov (2014)<sup>8</sup> | Cohort | Bulgaria | 84% | Median 56<br>Range 39–76 | NR | IIIB-IV | ADC, 56<br>SCC, 277<br>LCC, 14 | 20%<br>21%<br>21% |
| Du (2013)<sup>9</sup> | Cohort | China | 76% | Median 53<br>Range 31–75 | NR | IIIA-IV | ADC, 163<br>SCC, 95 | 61%<br>55% |
| Kim (2014)<sup>10</sup> | Cohort | Korea | 75% | Median 65<br>Range 20–84 | 68% | I-III | ADC, 107<br>SCC, 75 | 3%<br>16% |
| Michael (1999)<sup>11</sup> | Cohort | Canada | 65% | Median 63 | NR | LD-ED | SCLC, 46 | 33% |
| Nankano, T (1986)<sup>12</sup> | Cohort | Japan | NR | NR | NR | I-IV | ADC, 53<br>SCC, 56<br>LCC, 12<br>SCLC, 39 | 8%<br>22%<br>42%<br>5% |
| Olesen (1988)<sup>13</sup> | Cohort | Denmark | NR | NR | NR | NR | ADC, 18 | 28% |
| Pederson (2003)<sup>14</sup> | Cohort | Denmark | 80% | Median 63 | NR | I-IV | ADC, 13<br>SCC, 20<br>LCC, 7<br>SCLC, 11 | 54%<br>60%<br>57%<br>36% |
| Skorek (2018)<sup>15</sup> | Cohort | Poland | 64% | 67 (9) | 63% | I-IV | ADC, 128<br>SCC, 130<br>LCC, 16 | 9%<br>11%<br>13% |
| Tomita (2010)<sup>16</sup> | Cohort | Japan | 63% | 67<br>Range 26–86 | NR | I-III | ADC, 210 | 3% |
| Mean platelet count |  |  |  |  |  |  |  |  |
| Study | Study Design | Country | Males (%) | Mean age, years (SD) | Smoking history (%) | Clinical stage | Histological subtype, n | Platelet count (SD) |
| Biswas (2010)<sup>17</sup> | Cohort | India | 93% | 56<br>Range 30-76 | NR | I-III | ADC, 20<br>SCC, 28<br>LCC, 4<br>SCLC, 9 | 247 (25)<br>257 (20)<br>237 (28)<br>235 (19) |
| Demirkazik | Cohort | Turkey | 95% | 59 (SE: 2) | NR | IIIB-IV | ADC, 17 | 326 (SE: 28) |
| (2010)<sup>18</sup> |  |  |  |  |  |  | SCC, 38 | 340 (SE: 23) |
| Karagoz (2009)<sup>19</sup> | Case-Control | Turkey | 75% | Median 67<br>Range 39–92 | NR | LD-ED | SCLC, 17 | 297 (88) |
| Li, M (2019)<sup>20</sup> | Case-Control | China | 67% | 56 | 55% | I-III | ADC, 476 | 267 (68) |
| Nankano, K (2018)<sup>21</sup> | Case-Control | Japan | 77% | 71 (6) | 92% | IIB-IV | ADC, 11<br>SCLC, 10<br>SCLC, 5 | 298 (105)<br>328 (121)<br>291 (153) |
| Nankano, T (1986)<sup>12</sup> | Cohort | Japan | NR | NR | NR | I-IV | ADC, 53<br>SCC, 56<br>LCC, 12<br>SCLC, 39 | 265 (74)<br>274 (102)<br>353 (124)<br>249 (85) |
| Olesen (1988)<sup>13</sup> | Cohort | Denmark | NR | NR | NR | NR | ADC, 18 | 363 (116) |
| Sakin (2019)<sup>22</sup> | Cohort | Turkey | 80% | 59<br>Range 42–83 | 97% | I-III | SCLC, 90 | 303 (128) |
| Unsal (2004)<sup>23</sup> | Cohort | Turkey | 95% | 61<br>Range 36–74 | NR | IB-IV | ADC, 16<br>SCC, 22<br>SCLC, 20 | 356 (136)<br>410 (141)<br>400 (115) |
| Xu (2010)<sup>24</sup> | Case-Control | China | 66% | 57<br>Range 46–72 | NR | I-IV | ADC, 14<br>SCC, 18 | 262 (107)<br>197 (57) |
| Zhang (2015)<sup>25</sup> | Cohort | China | 69% | 60 (10) | NR | IA–IV | ADC, 308 | 247 (92) |
| Zhu (2014)<sup>26</sup> | Cohort | China | 67% | Median 56<br>Range 23–80 | 51% | IV | ADC, 246 | 276 (102) |

The search also returned an additional 18 studies with a further 6,917 lung cancer patients that did not fulfil the criteria for a meta-analysis, and are therefore discussed narratively. There was an inadequate number of studies per subtype for six of studies, seven studies defined thrombocytosis as a value other than 400 × 10 ^9^/L, and eleven reported median platelet count (some studies included more than one of these criteria). Eleven of these were assessed as ‘good’, one as ‘fair’, and six as ‘poor’ based on the Newcastle-Ottowa Scale. ^6^

### Meta-analysis of thrombocytosis proportions

We pooled the proportions of patients with thrombocytosis (defined as 400 × 10^9^ /L). ^7-16^ The overall pooled percentage of lung cancer patients with thrombocytosis was 27% (95% CI: 17 to 37%). By subtype, this proportion was 22% (95% CI: 7 to 41%) for ADC patients; 28% (95% CI: 15% to 43%) for SCC patients; 36% (95% CI: 13 to 62%) for LCC patients; and 30% (95% CI: 8 to 58%) for SCLC patients (Figure 2). There was little evidence of differences in thrombocytosis prevalence between these subtypes (p=0.76). The heterogeneity (I2 statistic) across subtypes was 97% for ADC, 92% for SCC, 61% for LCC, 86% for SCLC, and 93% across all studies. This was not affected by any of the sensitivity analyses.

**Figure 2:**
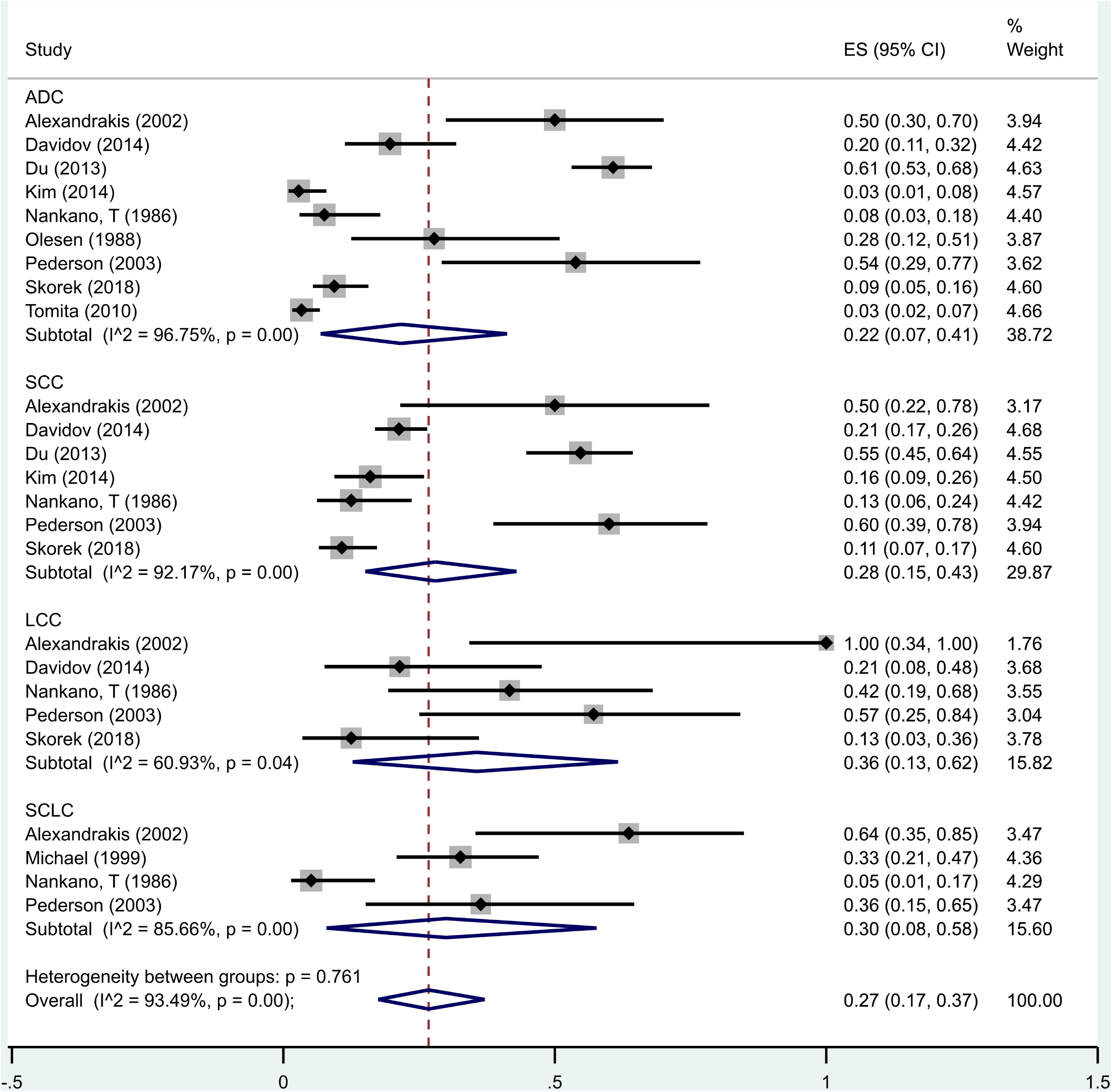
Random effects meta-analysis of studies that reported the proportion of patients with thrombocytosis. ^7-16^ Bars, 95% CI.

### Meta-analysis of mean platelet count

Of the 12 studies that reported a mean platelet count, ^12 13 17-26^ the pooled mean across all histological subtypes of lung cancer was 289 × 10 ^9^/L (95% CI: 268 to 311) (Figure 3). By subtype, the pooled mean platelet counts were 282 × 10 ^9^/L (95% CI: 259 to 306) for ADC; 297 × 10^9^ /L (95% CI: 238 to 356) for SCC; 290 × 10^9^ /L (95% CI: 176 to 404) for LCC; and 293 × 10^9^ /L (95% CI: 244 to 342) for SCLC. The I^2^ statistic for heterogeneity across studies was: 95% for ADC, 96% for SCC, 89% for LCC, 93% for SCLC, and 97% overall. There was little evidence of differences in mean platelet count across the histological subtypes of lung cancer (p=0.96). None of the sensitivity analyses changed the main results; however, the analysis restricted to advanced stage cancers reduced heterogeneity for the ADC and SCC subgroups to 46% and 0%, respectively. There were too few studies for LCC and SCLC to conduct a sensitivity analysis exclusively for advanced staged cancers.

**Figure 3:**
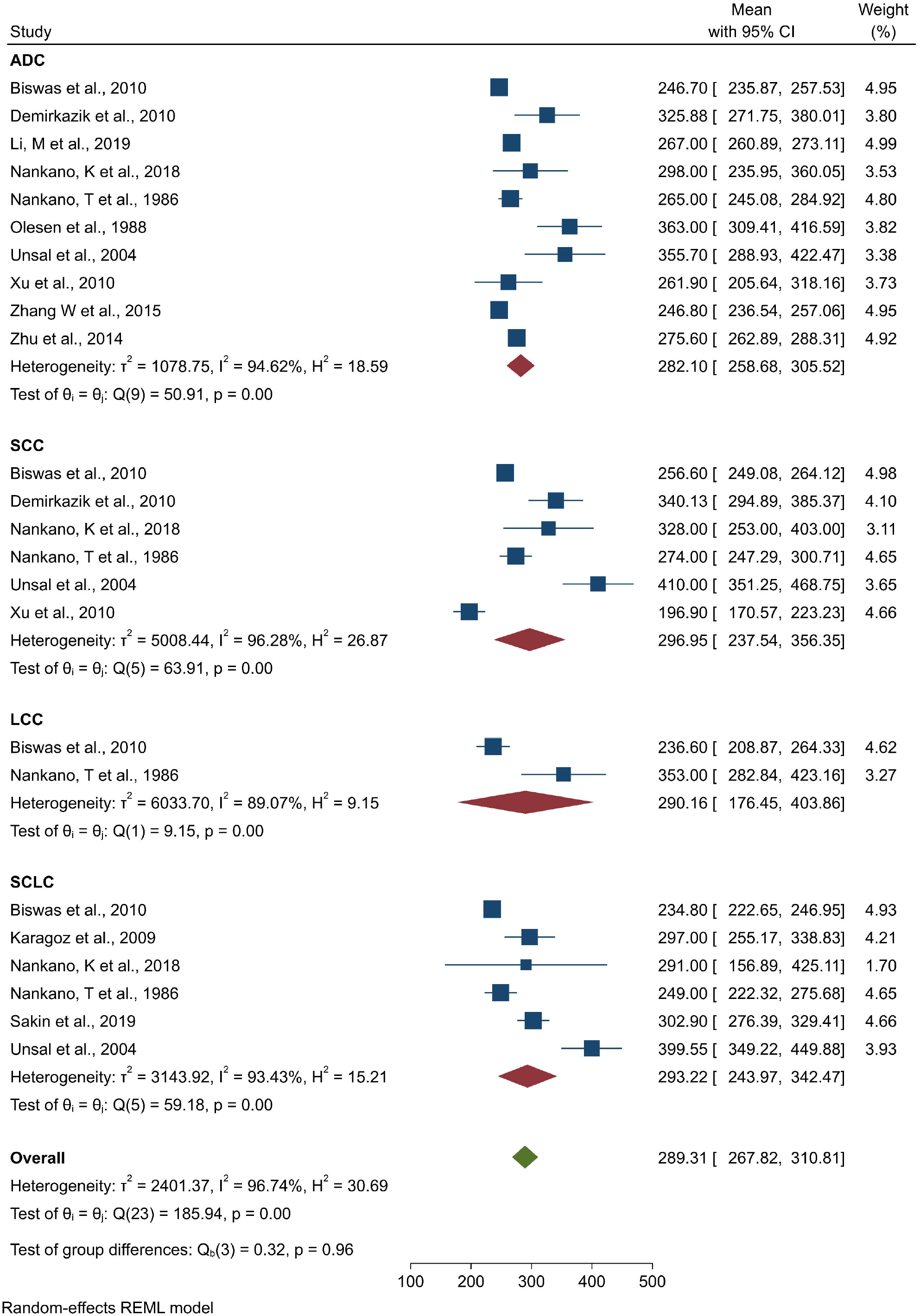
Random effects meta-analysis of studies that reported mean platelet count. ^12 13 17-26^ Bars, 95% CI.

### Median platelet counts and thrombocytosis proportions using different thresholds

There was also little evidence of differences across lung cancer subtypes in 11 studies, inclusive of 4,887, lung cancer patients that reported median platelet count values, ^27-37^ and seven studies of 2,795 lung cancer patients that used a different threshold to define thrombocytosis. ^29-31 37-40^

### Other subtypes of lung cancer

Six studies, totalling 841 patients, were returned that examined platelet levels across other subtypes of lung cancer. ^24 37-41^

#### Adenosquamous carcinoma

Two studies had available platelet counts or thrombocytosis proportions for patients with ADSq: Skorek and colleagues (2018)^15^ reported 1/11 (9%) patients with ADSq had thrombocytosis (400 × 10^9^ /L threshold), and Wang and colleagues (2017)^41^ reported a mean (SD) platelet count of 143 (65) across 134 patients.

#### ALK rearrangements

He and colleagues (2019)^42^ studied 344 patients with ALK-positive lung cancer and stratified patients’ platelet counts by percentage of ALK-positive tumour cells (15-49% and ≥50%), as detected by fluorescence *in situ* hybridisation. They reported 57% of patients with 15-49% of ALK-positive tumour cells and only 33% of patients with ≥50% ALK-positive tumour cells as having a platelet count of over 300 × 10^9^ / L blood. Yang and colleagues (2019)^43^ reported a median platelet count of 246 for 113 ALK-positive advanced NSCLC patients.

#### EGFR mutations

Two papers studied 112 ^44^ and 127 ^45^ advanced NSCLC patients, with either an *EGFR* exon 19 deletion or an *EGFR* exon 21 substitution mutation (L858R). They reported median platelet counts of 290^44^ and 232 × 10 ^9^/L, respectively.

## DISCUSSION

This is the first systematic review to compare platelet count across different subtypes of lung cancer. Data from 2,974 patients across 20 independent studies^7-26^ were synthesised and meta-analyses examined differences in mean platelet count and proportion of patients with thrombocytosis across ADC, SCC, LCC, and SCLC, finding no difference in mean platelet count or the percentage with thrombocytosis across the four subtypes. Similar results were observed in a further 18 studies of 6,917 additional participants that reported median platelet counts^27-37^ or proportion of patients with thrombocytosis using a lower threshold platelet count. ^29-31 37-40^ There were too few papers on adenosquamous, ^15 41^ *ALK-*positive, ^42 43^ and *EGFR*-positive ^44 45^ lung cancers to perform a meta-analysis, although the available evidence suggested thrombocytosis was not a prominent feature of any of these subtypes. Our results indicate that the relationship between pre-treatment thrombocytosis and lung cancer is a generic one, rather than being restricted to one or more cancer subtypes.

### Strengths and Limitations

This systematic review collated data from a very large number of lung cancer patients to analyse platelet count and proportion of thrombocytosis. We were meticulous in studying only pre-treatment platelet values: lung cancer therapies such as surgery, chemotherapy, and radiotherapy can affect platelet count. While most studies were assessed as ‘poor’ based on the Newcastle-Ottawa Scale, ^6^ all studies scored highly in the ‘selection’ domain. As the key data points extracted from each study were subtype diagnosis and platelet count, found in the patient characteristics rather than the results of the study, the ‘selection’ domain was the most important for the purpose of this review. Importantly, subtype diagnosis and platelet count were both obtained by robust, internationally recognised methods. The sensitivity analysis by study quality supported this. Furthermore, as this systematic review was concerned with patient characteristics (lung cancer subtype, platelet count) rather than the specific outcome of the included studies, any effect of publication bias should be small or absent.

There was a high level of heterogeneity throughout. To explore this, sensitivity analyses were conducted to analyse the effect of lung cancer stage, study quality and sample size. The sensitivity analysis that only included advanced lung stage cancers was the only one to reduce heterogeneity, perhaps because advanced stage cancers are more strongly associated with raised platelet count. ^2^ Another source of heterogeneity may be attributed to smoking status, as smoking is not only a risk factor for lung cancer but also elevates the platelet count. ^46^ Only a small number of studies reported smoking status but none differentiated smoking status in terms of platelet count and subtype: therefore, we were unable to analyse the effects of this.

### Comparison with the existing literature

We identified 27% (95% CI: 17 to 37%) of lung cancer patients to have pre-treatment thrombocytosis; this is much higher than the 1.5-2.2% found in the general (healthy) population ^3^ This is in keeping with previous studies: Hamilton and Peters (2005)^47^ reported that 26% of lung cancer patients had thrombocytosis and Bailey *et al* (2017)^3^ found 23% of males with undiagnosed lung cancer present in primary care with thrombocytosis 12 months before their diagnosis. We also reported an overall mean platelet count of 289 × 10^9^ /L (95% CI: 268 to 311) for lung cancer patients, again above the estimated mean platelet count of 258 × 10^9^ /L (SD: 62) for healthy individuals. ^48^ As these figures did not vary between lung cancer subtypes, we propose there is a generic relationship between lung cancer and a raised platelet count across the major lung cancer subtypes. This may reflect the lungs being the site of 50% of platelet biogenesis, ^49^ which could be stimulated by over-expression of cytokines and growth factors in the tumour microenvironment, irrespective of the tissue subtype. He *et al*., (2019)^42^ reported a reduced proportion of thrombocytosis in patients with ≥50% ALK-positive tumour cells, compared to patients with 15-49% ALK-positive tumour cells, which may be suggestive of a negative association between *ALK* translocations and thrombocytosis. As the only known study to report such a finding, it requires confirmation.

Platelets assist tumour metastasis in a number of ways. Tumour cells activate platelets upon dissemination, causing them to surround and protect the disseminated tumour cells from immune destruction and sheer stress to facilitate their haematological metastasis. ^50^ Furthermore, platelet-derived growth factors promote endothelial permeability to expedite transendothelial migration for tumour extravasation in distant tissues and organs. ^51^ Platelet-derived growth factors also stimulate tumour cell proliferation in order to establish and sustain secondary tumour growth at these distant sites. Of note, the platelet-derived growth factor pathway is regulated by the tumour suppressor p53, encoded by the *TP53* gene, which is commonly mutated in lung cancer patients. ^52^ Unfortunately, there is no existing literature regarding platelet count in lung cancer patients with *TP53* mutations.

Whether thrombocytosis is due to the development of lung cancer or whether lung cancer development is enhanced by thrombocytosis is as yet unknown, and it may be truly bidirectional. Thrombocytosis can be a reactive condition that develops secondary to other disease, including lung cancer. ^53^ However, a recent Mendelian randomisation analysis suggested a causal relationship between raised platelet count and increased lung cancer risk. ^54^

### Clinical implications

These findings will be of interest to clinicians involved in diagnosing and estimating prognosis in lung cancer. Platelet count has long been used as a marker of prognosis in several cancer types; these results show that an elevated platelet count is no more strongly indicative of one lung cancer subtype over any other, and that lung cancer should be considered in a patient with an otherwise unexplained raised platelet count.

## CONCLUSION

This systematic review and meta-analysis is the first to assess the relationship between platelet count and the subtypes of lung cancer. We report a 27% prevalence (95% CI: 17% to 37%) of thrombocytosis across lung cancer patients and a mean platelet count of 289 × 109/L (95% CI: 268 to 311). We report no evidence of differences in platelet levels across the major subtypes of lung cancer and therefore suggest that a thrombocytosis is likely to be a generic feature across all lung cancer subtypes.

## Data Availability

Data is available by contacting Melissa Barlow (corresponding author)

